# Trends in Antibody Titers after SARS-CoV-2 Vaccination - Insights from a Self-Paid Tests at a General Internal Medicine Clinic

**DOI:** 10.1101/2023.03.03.23286284

**Authors:** Hiroshi Kusunoki, Kazumi Ekawa, Masakazu Ekawa, Nozomi Kato, Keita Yamasaki, Masaharu Motone, Hideo Shimizu

## Abstract

**Background:** The rise in antibody titers against the novel coronavirus (SARS-CoV-2), and its duration is considered an important indicator for confirming the efficacy of the COVID-19 vaccine, and self-paid tests of antibody titer are conducted at many facilities nationwide.

**Methods:** The relationship between the number of days after the second and third dose of vaccines, age, and antibody titer was determined from the medical records of general internal medicine clinics that conducted self-paid testing of SARS-CoV-2 antibody titer using Elecsys Anti-SARS-CoV-2 S (Roche Diagnostics), as well as the relationship between the number of days after two or more dose of vaccines and antibody titer. We also examined antibody titers in cases of spontaneous infection with SARS-CoV-2 after two or more doses of the vaccine.

**Results:** Age and antibody titer were negatively correlated in 86 subjects whose antibody titer was measured within 1 month after the second vaccination with the COVID-19 vaccine. In 31 subjects whose antibody titer was measured within one month after the third dose of vaccine, age and antibody titer showed a negative correlation (p<0.01). The mean antibody titer after the third vaccination was 20704.9±16820.7 U/mL, more than 10 times the mean antibody titer after the second dose of vaccine of 1601.9±1297.9 U/mL. There were also cases of infection after the third or fourth dose of vaccine, with antibody titers in the tens of thousands after infection, but they still received further booster vaccinations after infection.

**Discussion:** SARS-CoV-2 antibody titers measured within 1 month of the second or third dose of vaccine showed a negative correlation with age, and antibody titers also showed a negative correlation trend with the number of days after the second dose of vaccine. It is considered that many people in Japan received further booster vaccinations after spontaneous infection, even though they already had antibody titers in the tens of thousands U/mL by “hybrid immunity” after spontaneous infection following two or more dose of vaccine.

**Conclusion:** After measuring SARS-CoV-2 antibody titers, booster vaccination of those with low antibody titers should be done on a priority basis.

## Introduction

The COVID-19 vaccine has been available in Japan since February 2021 for healthcare workers and since April 2021 for the elderly citizens. As of December 10, 2021, 77.3% of the total population had completed two vaccination doses. The third dose of vaccinations also began in December 2021 for healthcare workers, and in early 2022, vaccinations began in earnest for the general population, starting with the elderly individuals.

Furthermore, from the end of May 2022, the fourth dose of vaccine was given after a gap of at least 5 months after the third dose, for those aged 60 years or older, those aged 18 years or older with underlying medical conditions, and other patients diagnosed by physicians to be at high risk of severe COVID-19.

Moreover, the vaccination interval for Pfizer and Moderna vaccines has been shortened from 5 months to 3 months starting in October 2022. Accordingly, the fifth dose of vaccine has been started. Thus, it can be said that COVID-19 vaccination has been promoted under the government’s initiative in Japan.

Severe acute respiratory syndrome coronavirus 2 (SARS-CoV-2) antibody titers level and duration of elevated levels are considered important indicators of the efficacy of COVID-19 vaccines. Many facilities in Japan conduct self-paid tests for SARS-CoV-2 antibody titers.

“Hybrid immunity” of vaccination plus spontaneous infection brought about by breakthrough infection after vaccination is gaining worldwide attention as COVID-19 vaccination becomes more widespread. The strength of “hybrid immunity” with the rise in, and duration of, antibody titers against SARS-CoV-2 after post-vaccination infection are considered important indicators for confirming the efficacy of COVID-19 vaccines. It has been reported that breakthrough infection after vaccination markedly increases antibody titers ^1^.

Recently we had reported that when patients are spontaneously infected with SARS-CoV-2 after the second or more dose of vaccine, antibody titers increased to more than 40,000 AU/mL and remained high for several months ^2^.

Participants in our previous study included many employees of Osaka Dental University Hospital and students of Osaka Dental University, many of whom are healthcare workers at high risk of infection with COVID-19 on a daily basis. In addition, the participants were relatively young people living in metropolitan areas, and may not reflect the general population. Elderly people may not be able to produce antibodies due to sufficient immune response by immune memory of B cells, even in the case of breakthrough infection after two or more doses of vaccine. It is also unclear whether other assay systems for SARS-CoV-2 antibody would yield similar results.

In this study, we examined changes in SARS-CoV-2 antibody titers in outpatients who had their SARS-CoV-2 antibody titers measured by self-paid test after vaccination with the COVID-19 vaccine in general internal medicine clinics in rural areas.

## Materials and methods

This was a single-center retrospective study. We conducted a longitudinal observational study involving 133 outpatients (47 males and 86 females, mean age 65.6±19.1years) at the Ekawa Medical Clinic, Arida City, Wakayama, Japan, between June 2021 to September 2022. The study protocols were approved by the ethics committee of Osaka Dental University Hospital (2022-10). Written informed consent was obtained from all participants. The titers of IgG-type SARS-CoV-2 antibody titers SARS-CoV-2 anti-receptor-binding domain IgG antibodies were measured in serum samples.

First, we showed typical antibody titers in married couples who were vaccinated with COVID-19 vaccines, after the third and fourth doses, respectively..

Second, the correlation between antibody titer, age, and the number of days after the second dose of the vaccine was examined in 86 subjects (36 males and 50 females) with no history of COVID-19 infection, and the date of vaccination was known. The same examination was also conducted on 31 subjects (11 males and 20 females) with no history of COVID-19 infection after the third dose of the vaccine, and the date of vaccination was known. The post-infection antibody titers of subjects infected with COVID-19 after two or more doses of the COVID-19 vaccine were also examined.

### Serology assays

We used the electrochemiluminescence immunoassay system Elecsys Anti-SARS-CoV-2 S (Roche Diagnostics, Mannheim, Germany), to detect IgG antibodies to the receptor-binding domain of the S subunit of the SARS-CoV-2 spike protein, according to the manufacturer’s instructions. The cut-off value set by the manufacturer was 0.8 U/mL. A strong correlation was observed between the measurements of Abbott Architect SARS-CoV-2 IgG II Quant (Abbott Laboratories, Illinois, USA), the assay system for the antibody we used in our previous report ^2^, and those of Elecsys Anti-SARS-CoV-2 S ^3^.

### Statistical analysis

The results are expressed as mean ± standard deviation (SD). Pearson’s product-moment correlation coefficient was used to assess the associations between SARS-CoV-2 anti-receptor binding domain IgG antibody titers, age, and number of days after vaccination. For data analysis, the JMP 13.1 software was used for data analysis. Statistical significance was set at p < 0.05.

## Results

Figure 1A shows a couple in their 60s who had received the third dose of COVID-19 vaccine. They had both completed the second dose by June 2021, but their antibody titers then decayed, with the wife receiving her third dose in March 2022 and the husband in May 2022. Both of them had much higher antibody titers than after the second dose, up to 10,000U/mL. The titers then decayed again over the next two to three months, but still remained much higher than the peak after the second dose of vaccine.

**Figure 1A.**
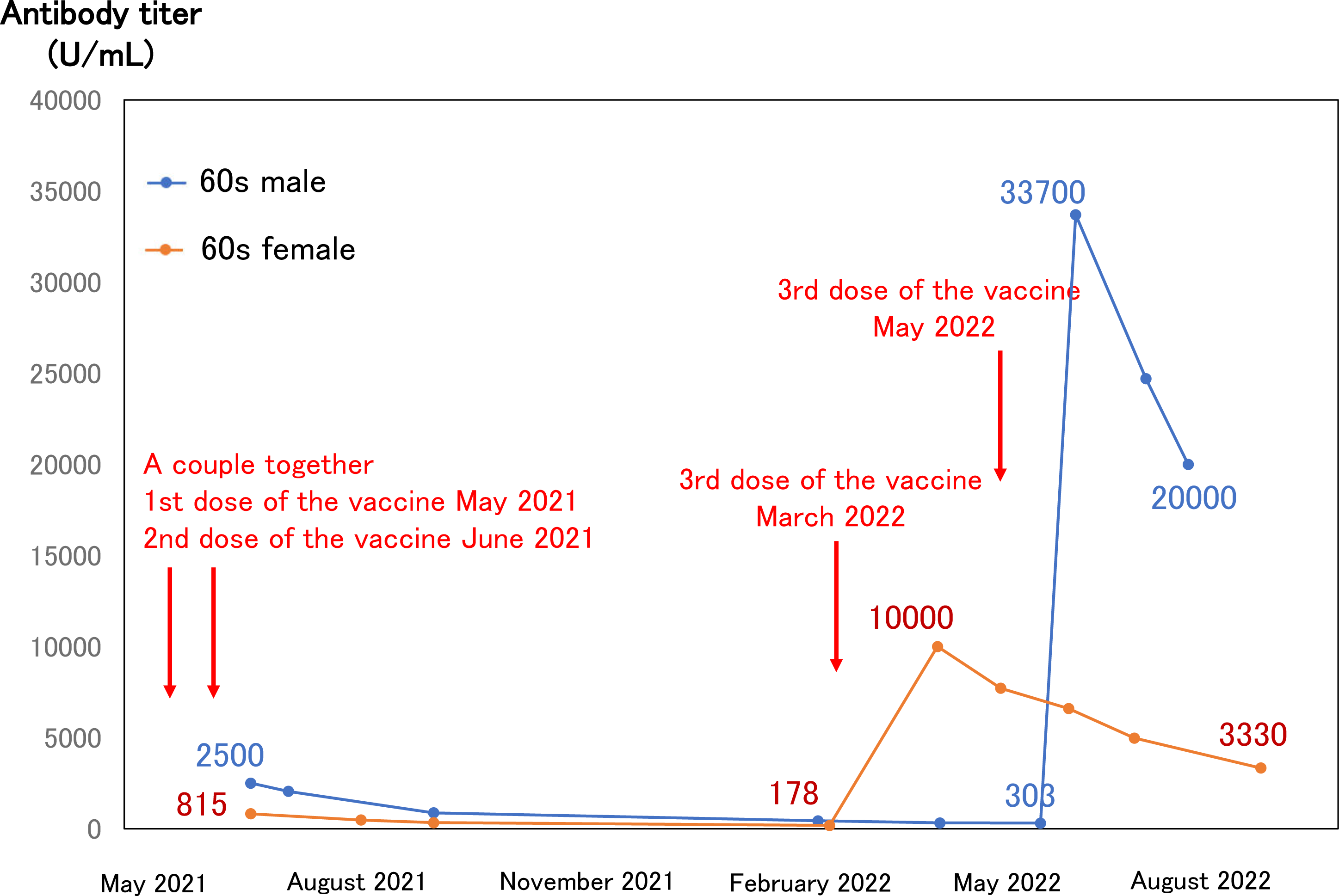
An example of a couple vaccinated up to the third dose.

**Figure 1B.**
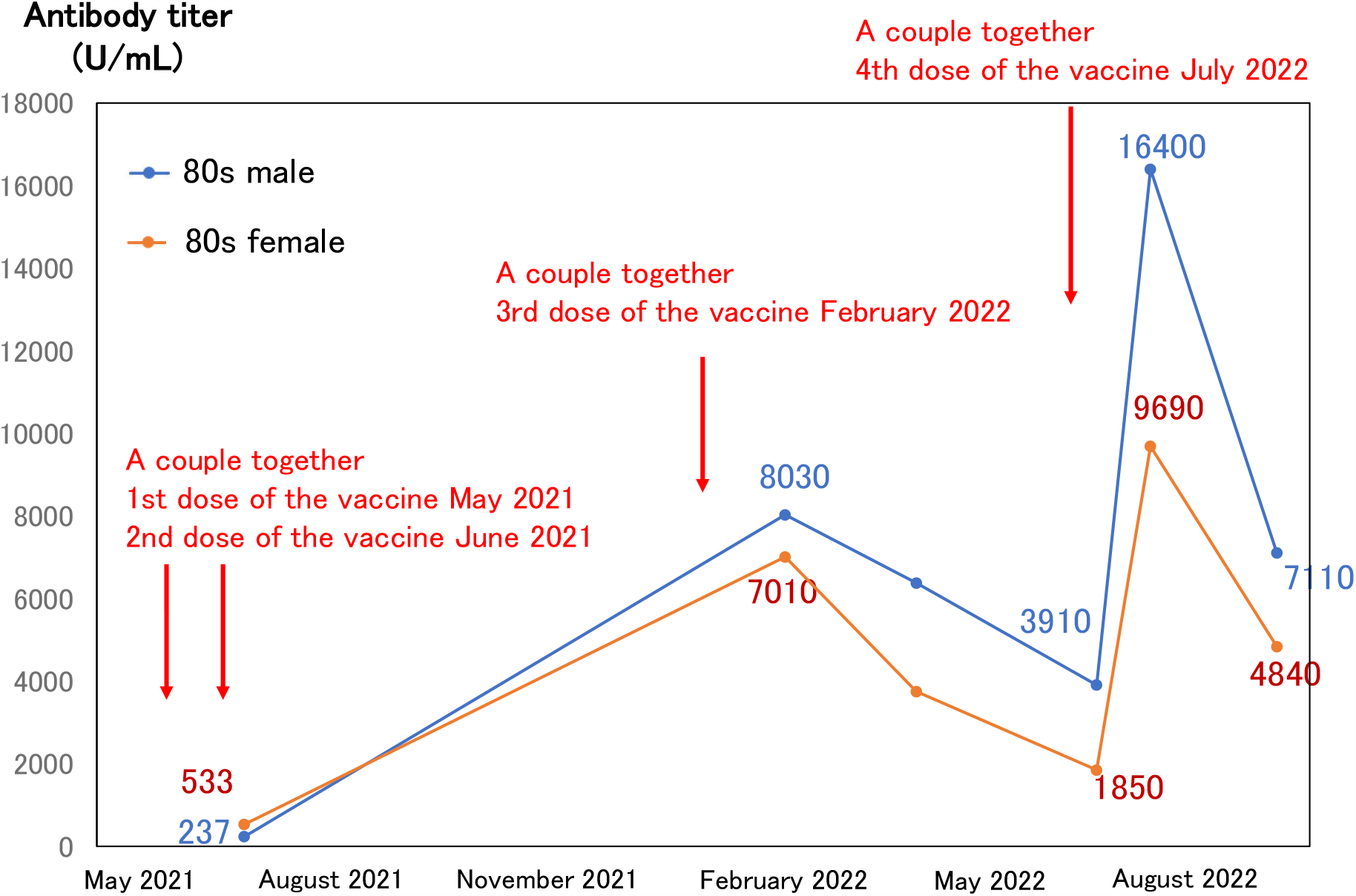
An example of a couple vaccinated up to the fourth dose.

Figure 1B shows an example of a late-elderly couple in their 80s who had received the fourth dose of vaccine. The first two doses of vaccines were administered by June 2021. The third dose of vaccine was given in February 2022, and the antibody titer rose to 7,000-8,000 U/mL immediately. This then dropped over the next 5 months or so, and when the fourth dose of vaccine was given in July 2022, it rose again, but then waned over the next 2 months.

Antibody titers were measured in 86 subjects (36 males and 50 females) after the second dose of the vaccine. These subjects had no history of COVID-19 infection at the time of antibody titer measurement. The mean antibody titer was 1601.9±1297.9U/mL (Figure 2A).

**Figure 2A.**
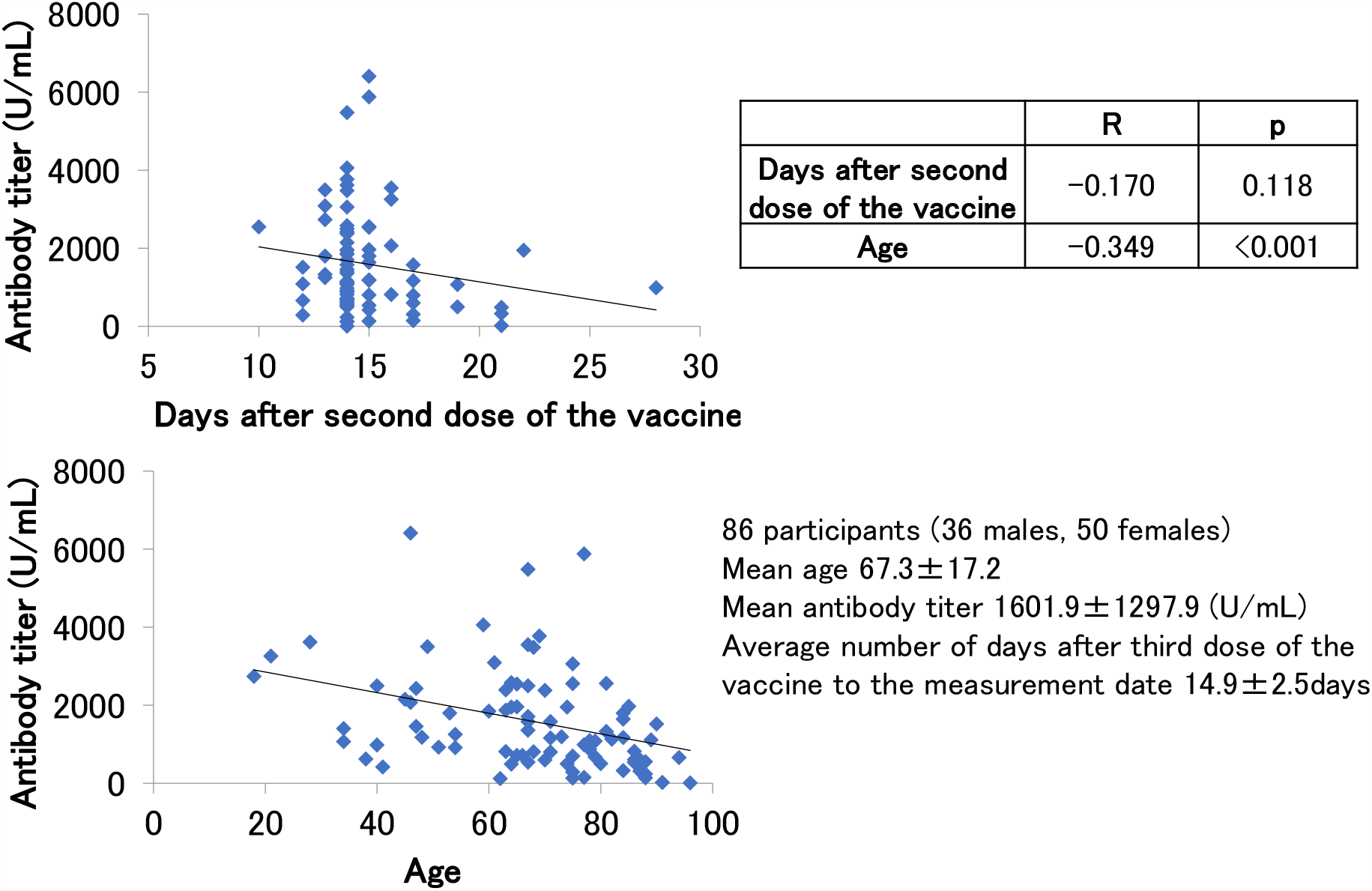
Days after 2nd dose, age and antibody titer.

**Figure 2B.**
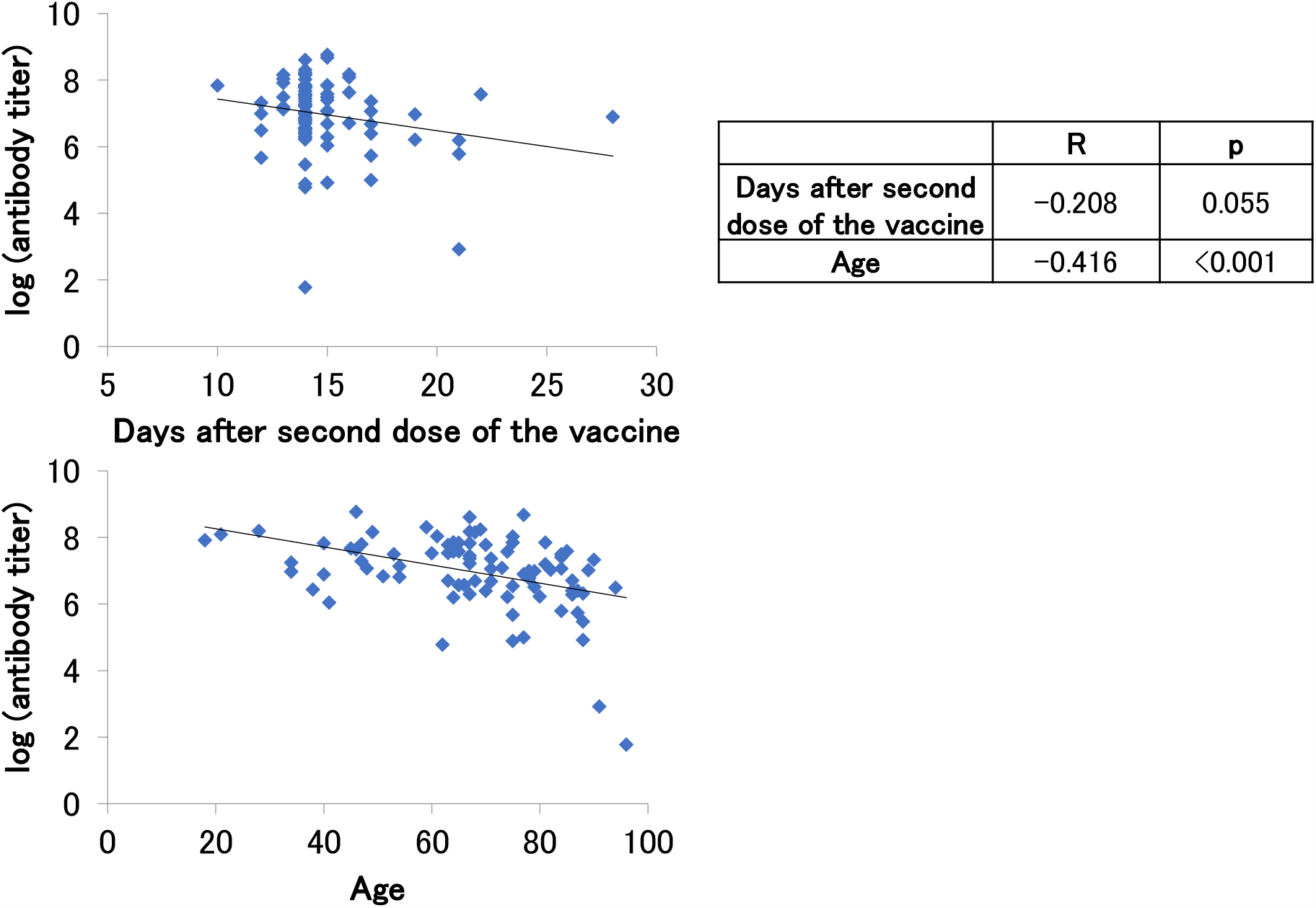
Days after 2nd dose, age and log (antibody titer)

Since all cases were tested at private clinics at their own expense, most people came to see if they had enough antibody titers after vaccination, so antibody titers were measured within one month after vaccination. On an average, antibody titers were measured about two weeks after vaccination.

We did not find significant correlation between antibody titer and number of days after the second dose of the vaccine in a period of time as short as one month, but a trend for antibody titers to decrease was observed (p=0.118). On the other hand, there was a significant negative correlation between age and antibody titer (p<0.001).

Another negative correlation trend was observed between log-transformed antibody titer and days after second dose of the vaccine, although this was not significant (p=0.055). A significant negative correlation was also observed between log-transformed antibody titer and age (p<0.001) (Figure 2B).

Antibody titers were measured in 31 subjects (11 males and 20 females) with no history of COVID-19 infection at the time of antibody titer measurement after the third booster dose of the vaccine (Figure 3A). The average antibody titer after the third dose of the vaccine was 20704.9± 16820.7 U/mL, which was certainly boosted from that after the second dose of the vaccine.

**Figure 3A.**
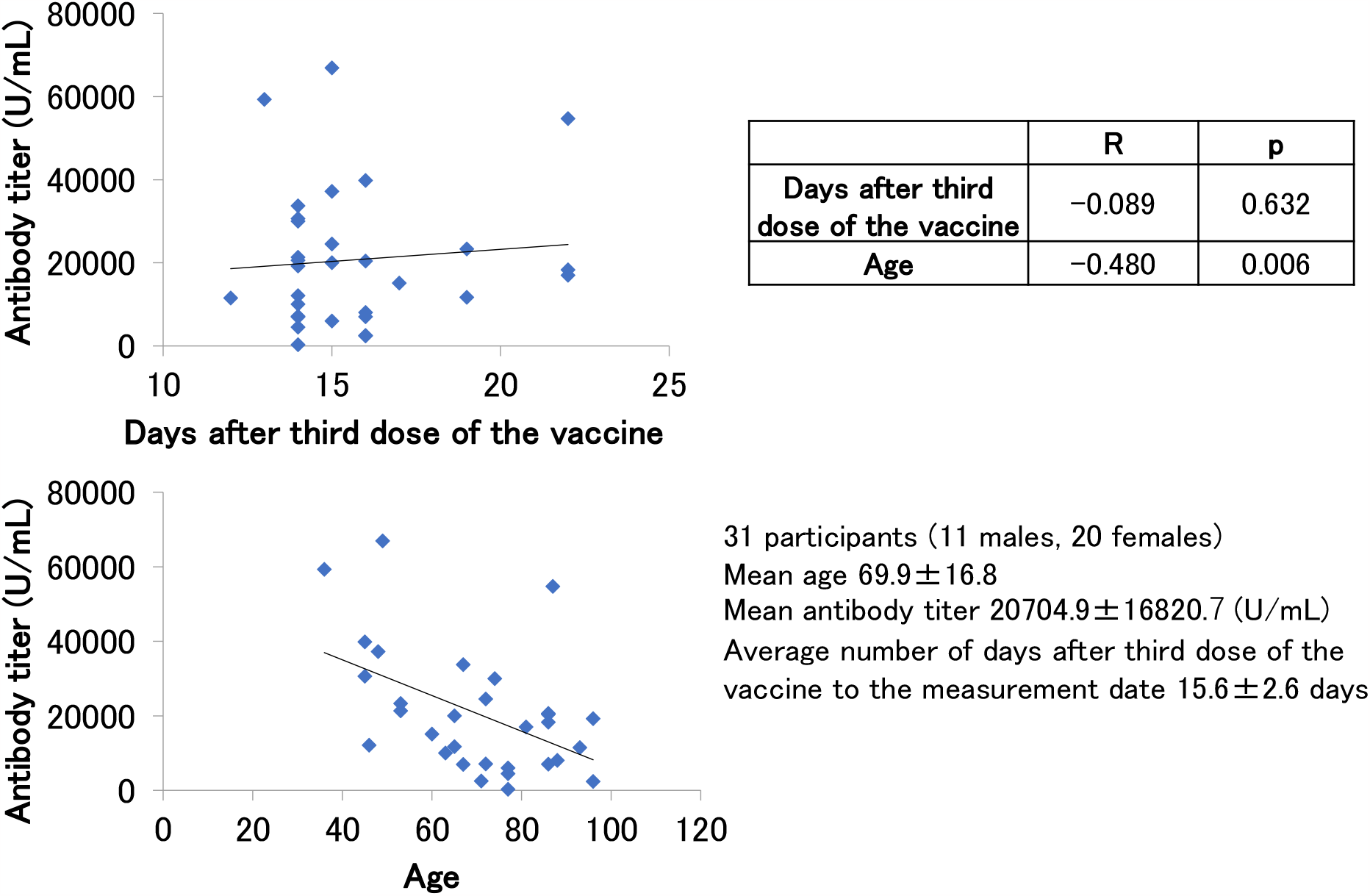
Days after 3rd dose, age and antibody titer.

**Figure 3B.**
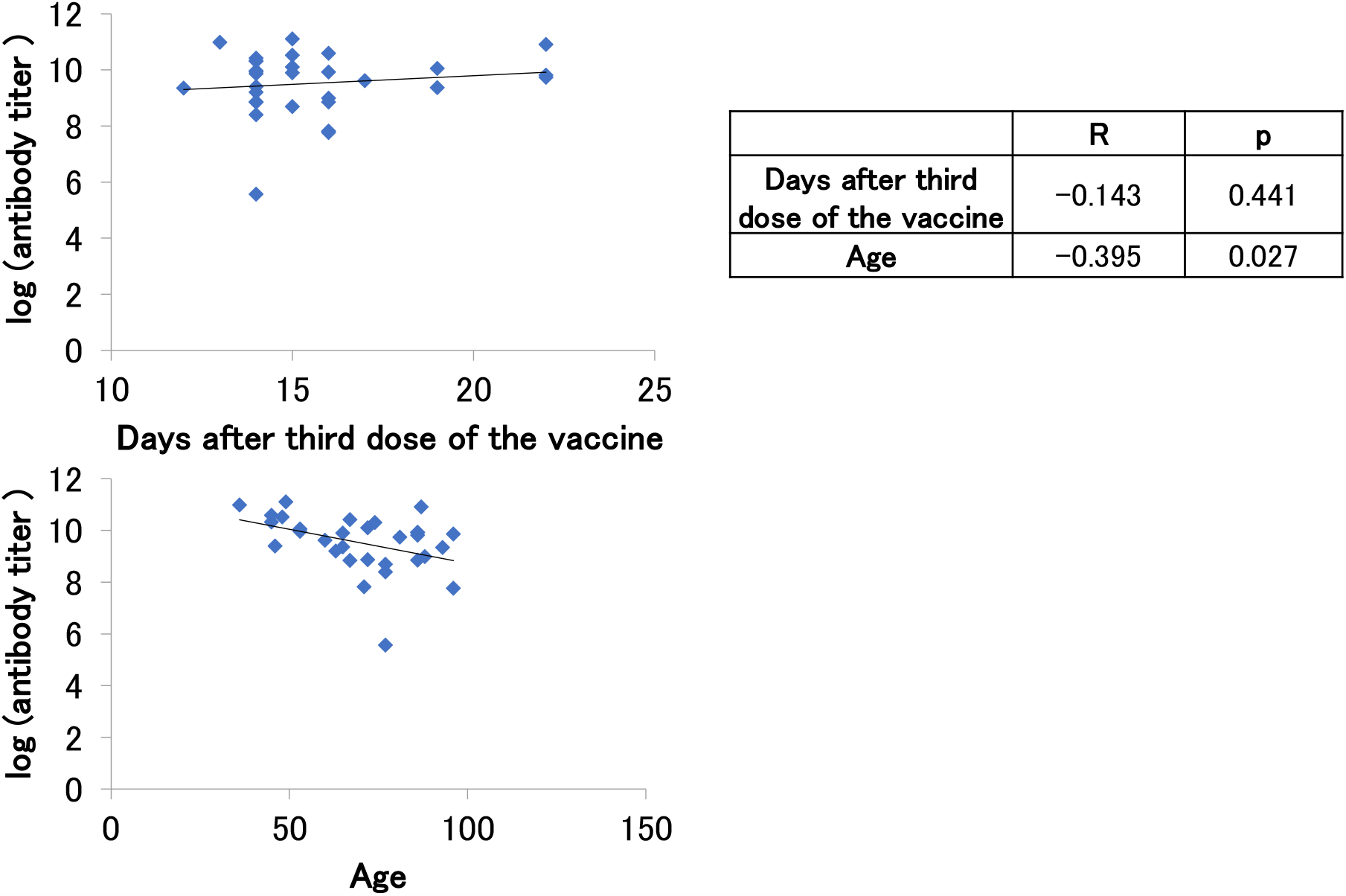
Days after 3rd dose, age and log (antibody titer)

There was no significant correlation between antibody titer after the third dose of vaccine and the number of days after third dose of the vaccine (p=0.632) (Figure 3A). No significant correlation was observed between the log-transformed antibody titer after the third dose of vaccine and the number of days post-vaccination (p=0.414) (Figure 3B). On the other hand, as with the antibody titer after the second dose of vaccine, both the antibody titer after the third dose of vaccine and the log-transformed antibody titer after the third dose of vaccine showed a significant negative correlation with age (Figure 3A, B).

Table 1A shows two cases of spontaneous infection after the second dose of vaccine. A 40s male became infected about six months after the second dose of vaccine, and his antibody titer was over 100,000U/mL after infection. In a 50s female, the antibody titer had risen to 26,600U/mL after infection, but she had received her third vaccination a few days after the antibody titer was measured.

**Table 1.**

| Case | 1st vaccination date<br>2nd vaccination date | Date of infection | Days from 2nd<br>vaccination to<br>infection | Post-infection<br>antibody test date | Post-infection antibody<br>titer (U/mL) | Number of days from infection to<br>antibody titer measurement | 3rd vaccination date |
| --- | --- | --- | --- | --- | --- | --- | --- |
| 40s M | August 2021<br>September 2021 | March 2022 | 183 | April 2022 | 113000 | 37 |  |
| 50s F | unknown<br>unknown | January 2022 |  | March 2022 | 26600 | 48 | March 2022 |

B. Participants who got infected after the third dose of vaccine
| Case | 1st vaccination date<br>2nd vaccination date<br>3rd vaccination date | Date of infection | Days from 3rd<br>vaccination to<br>infection | Post-infection<br>antibody test date | Post-infection antibody<br>titer (U/mL) | Number of days from infection to<br>antibody titer measurement | 4th vaccination date |
| --- | --- | --- | --- | --- | --- | --- | --- |
| 60s F | June 2021<br>July 2021<br>February 2022 | March 2022 | 36 | April 2022 | 20500 | 14 | August 2022 |
| 70s F | June 2021<br>July 2021<br>March 2022 | July 2022 | 118 | July 2022 | 12200 | 11 |  |
| 50s F | unknown<br>unknown<br>March 2022 | February 2022 | 160 | September 2022 | 59600 | 26 |  |
| 70s M | June 2021<br>July 2021<br>March 2022 | August 2022 | 156 | August 2022 | 41000 | 18 | August 2022 |
| **70s F** | July 2021<br>July 2021<br>March 2022 | August 2022 | 153 | August 2022 | 9840 | 16 | August 2022 |
| **70s F** | unknown<br>unknown<br>February 2022 | March 2022 | 31 | April 2022 | 63400 | 45 | August 2022 |

C. Participants who got infected after the fourth dose of vaccine
| **Case** | **1st vaccination date**<br>**2nd vaccination date**<br>**3rd vaccination date**<br>**4th vaccination date** | **Date of infection** | **Days from 4th**<br>**vaccination to infection** | **Post-infection antibody**<br>**test date** | **Post-infection antibody titer**<br>**(U/mL)** | **Number of days from**<br>**infection to antibody**<br>**titer measurement** |
| --- | --- | --- | --- | --- | --- | --- |
| **80s F** | unknown<br>unknown<br>February 2022<br>July 2022 | August 2022 | 44 | September 2022 | 122000 | 23 |

Table 1B shows six cases of spontaneous infection after the third dose of vaccine. Although some of the cases had their antibody titers measured nearly six months after infection, they all maintained high levels of antibody titers. However, despite maintaining high levels of antibody titers, some of them had received the fourth dose of vaccine.

Table 1C shows a case of an elderly woman who was spontaneously infected after the fourth dose of vaccine. This case received the fourth dose of vaccine in July 2022, but she became spontaneously infected in August, and her antibody titer rose to about 120,000U/mL after infection. Thus, even elderly people seem to show a marked increase in antibody titer after infection.

## Discussion

The present study, showed that SARS-CoV-2 antibody titers were boosted by the third or fourth dose of vaccine, but then declined over time (Figure 1A, B). As observed in this study, immunity acquired by booster vaccination alone is known to fade faster ^4^.

Among the 86 participants whose antibody titers were measured within one month after the second dose of vaccine, age and antibody titer showed a significant negative correlation. A negative correlation trend between the number of days after the second dose of vaccine and antibody titer was observed, but was not significant (Figure 2A, B).

Similarly, in the 31 subjects whose antibody titers were measured within one month after the third dose of vaccine, age and antibody titer showed a negative correlation (Figure 3A, B). It has previously been reported that antibody titers after vaccination tend to decrease with increasing age ^5-8^.

The negative correlation between antibody titer after the second or third dose of vaccine and age in this study is consistent with previous reports. The average antibody titer after the third dose of vaccine was more than 10 times higher than that after the second dose of vaccine. In the 1-month post-vaccination follow-up period, no correlation was found between the number of days after the third dose of vaccine and antibody titer. It was hypothesized that third doses of vaccine would provide stronger immunity and less attenuation of antibody titers than after previous doses of vaccine.

Researchers in Israel studied more than 10,000 health-care workers who had not previously been infected; all of them received either three or four doses of the vaccine. That study showed that up to the second dose of vaccine, antibody titers tended to drop, but after the third dose of vaccine, antibody titers were maintained to some extent even after six months9. The result that antibody titers obtained after the third dose of vaccine are less attenuated than those obtained after the second dose of vaccine is consistent with our findings.

On the other hand, in the Israeli study^9^, the levels of antibody titers after the third and fourth doses of vaccine did not change much, with the antibody response after the fourth dose of vaccine peaking at about four weeks, declining to the same level as before the fourth dose of vaccine at 13 weeks, and then stabilizing^9^. These results suggest that the fourth dose of vaccine may be less significant in terms of maintenance of antibody titers. The authors also found that the fourth dose’s efficacy against infection fell rapidly. In fact, after four months, the fourth dose was no better than three doses at preventing infection.

A study of derived from the national database in Portugal showed that hybrid immunity, the result of both vaccination and a spontaneous infection by COVID-19, could provide partial protection against reinfection for at least eight months ^10^. A survey of health care workers in Sweden suggested that those with hybrid immunity have high levels of mucosal antibodies with strong protective effects against infection^11^. It also offers greater than 95% protection against severe disease or hospitalization for between six months and a year after an infection or vaccination, according to estimates from a meta-analysis ^12^.

In the present study, we showed that post-infection antibody titers were markedly elevated in post-vaccination breakthrough infection, even in a completely different population and using a different measurement system, from our previous study, which also showed similar results. At the same time, there were scattered cases in which even those with sufficiently elevated antibody titers due to breakthrough infection received additional doses of vaccine. It is debatable whether additional vaccinations should be administered, when a patient is infected and antibody titers have increased markedly.

Since the Omicron strain was found to cause low disease severity, the Japanese government has reviewed the status of COVID-19 under the Infectious Disease Control Law and plans to lower it to Category 5, the same as influenza infections, starting in May 2023. In Japan, COVID-19 vaccination has been recommended as frequently as possible in each epidemic surge, but in accordance with this change, there is a proposal to reduce the vaccination to once a year.

As shown in this study, those who were vaccinated three or more times had strong immunity. In Japan, most cases of SARS-CoV-2 infection occurred after two or more doses of vaccine. In such cases, as shown in our previous report^2^, the post-infection antibody titer increased markedly, and the high antibody titer may have continued for a long period of time (more than six months). It is necessary to clarify the duration of high antibody titers after breakthrough infection, whether they last for 6 months to 1 year. In cases where antibody titers were markedly increased by hybrid immunity of vaccination plus spontaneous infection, the incidence and severity of the COVID-19 were lower, thus providing strong evidence that frequent additional vaccinations are not necessary, at least for recently infected persons, and that annual vaccinations are sufficient.

In conclusion, as we mentioned in the previous reports^2^, an increase in antibody titers against SARS-CoV-2 and the duration for which it remains high are considered important indicators of the efficacy of novel coronavirus vaccines. Although it is desirable to measure antibody titers and prioritize those with low antibody titers for booster vaccination, rather than blindly recommending booster vaccination to the entire population, it may be difficult to see how the financial, personnel, time, and educational costs can be afforded. As this was a small single-center retrospective study, larger-scale longitudinal studies of antibody titers are warranted.

## Data Availability

All data produced are available online at

